# Sequential Vaccination for Containing Epidemics

**DOI:** 10.1101/2020.04.13.20060269

**Authors:** Guy Tennenholtz, Constantine Caramanis, Shie Mannor

## Abstract

The dynamics of infectious diseases spread is crucial in determining their risk and offering ways to contain them. We study sequential vaccination of individuals in networks, where there is a limit on the number of individuals that can be vaccinated every day. Effective allocation of vaccine will play a critical role in preventing the spread and reducing the effects of a future pandemic. We derive methods for calculating upper and lower bounds of the expected number of infected individuals, as well as provide estimates on the number of vaccinations that is needed for containment. We calculate these explicitly on trees, d-dimensional grids, and Erdős Rényi graphs. Finally, we construct a time-dependent budget allocation strategy and demonstrate its superiority over constant budget allocation on real networks following first acquaintance vaccination. Our results provide a principled approach to assess the needed vaccination rate given the social graph topology.

## 1. Introduction

Consider an outbreak of a contagious disease such as the novel Corona virus or the seasonal flu. Flu outbreaks happen every year and vary in severity, depending in part on what type of virus is spreading. The influenza or flu pandemic of 1918 was the deadliest in modern history, infecting over 500 million people worldwide and killing 20-50 million victims. Between the years 2014-2016 West Africa experienced the largest outbreak of Ebola in history, with multiple countries affected. A total of 11,310 deaths were recorded in Guinea, Liberia, and Sierra Leone. Beginning in December 2019, COVID-19, a new coronavirus, began appearing in China. COVID-19 was declared a pandemic in March 2020, and by April 2020 over 1 million people around the globe had been infected by it. With such pandemics, global connectedness may trigger a cascade of infections. The outbreaks of Ebola and COVID-19 echo scenarios where long-range routes of transmission, most prominently international air routes - can allow the deadliest viral strains to outrun their own extinction, and in the process kill vastly more victims than they would have otherwise. The most dangerous pathogens leave their hosts alive long enough to spread infection.

Although the overarching aim is to vaccinate all persons who choose to be vaccinated, prior to the peak of disease, the vaccine supply to meet this goal may be insufficient early in a pandemic. Supplies for vaccinating the vast population may be scarce as opposed to the contagion speed. For example, the vaccination may use antibodies that are extracted from recovered patients that are scarce and expensive to produce, or the vaccination may hold potential risks for some of the population. We consider immunization strategies that eliminate epidemic threats by vaccinating parts of the population. Infections may propagate quickly and be discovered only at late stages of propagation. Vaccination policies define rules for identifying individuals that should be made immune to the spreading epidemic. Vaccinating such individuals may stop the spread of the epidemic in what is sometimes referred to as “herd immunity”. In this paper we study sequential vaccination policies that use full information on the infectious network’s state and topology, under vaccination budget constraints.

The analysis of epidemic spreading in networks has produced results of practical importance, but only recently the study of epidemic models under dynamic control has begun. The control of epidemics has been extensively studied for the past two decades. The dynamic allocation of cure has been studied in [1, 2, 3, 4, 5]. Studies of vaccine allocation in [6, 7, 8, 9, 10, 11, 12]. An optimization strategy for optimal vaccine allocation is proposed in [7]. Their model assumes slightly modifiable infection rates and a cost function based on a mean field approach. Information-driven vaccination is studied in [9], showing the spread of the information will promote people to take preventive measures and consequently suppress the epidemic spreading. It was also proposed to only vaccinate those individuals with the most unvaccinated contacts [10]. An acquaintance immunization policy was proposed in [6], where it was shown that such a policy is efficient in networks with broad-degree distribution. A different approach [12], considers minimizing the social cost of an epidemic. The above studies’ analysis is based on epidemic thresholds and mean-field approximations of the evolution process. In contrast to these, this paper studies the transient, short-term behaviour of the spreading epidemic.

Previous research on immunization policies modeled the problem either deterministically or approximately, considering long-term effects alone. Defending against an attack of a virus must take into account its transient behavior, fast propagation, as well as natural budget constraints (e.g., limited rate in which vaccines can be allocated). It is thus vital to find on-line policies which acknowledge short-term effects and the defender’s limited protection capabilities.

An assumption we make in the analysis is that the network structure is *known*. That is, it is known when and how the nodes interact. For spreading pandemics such as the COVID-19, this assumption is justified because of two reasons. First, interaction maps using cellular phones (either through an app, or using information from the carrier) are available or can be made available in most countries. Second, for the conclusions of this paper to hold, a simulated social interaction graph that reflects the social interactions suffices to obtain credible results. Alleviating this assumption is possible, by, e.g., considering partial knowledge of the network structure, but we believe the conclusions of this paper still hold even if this assumption is relieved.

We start by developing the mathematical machinery needed to characterize the spreading epidemic. We then consider criteria for containing an epidemic by constructing upper and lower estimates on the vaccination budget needed to contain it. Finally we propose an algorithm for state-dependent budget allocation, based on estimated local growth rate behavior of the network. We show that this strategy achieves better containment on two real world networks, when compared to a constant budget strategy which consumes an equal global budget. We conclude the paper with insights on the COVID-19 pandemic.

## 2. Model

This section defines the model of our problem and the criteria we wish to optimize. We will distinguish between two different criteria. The first, and most natural criterion, considers minimizing the number of infected individuals at the end of the infection process. As this criterion is hard to optimize generally [13, 14], we consider an alternative criterion, which is ultimately easier to evaluate. The containment criterion asks the following question: What is the minimal budget needed to ensure an infection won’t reach a certain size? This question can be answered immediately if the first criterion is minimized, whereas knowing the latter does not give us the former. Strictly speaking, this question may be more feasible to answer, and is thus the main focus of our work. In regard to the optimality criterion, we provide upper and lower bounds on the number of infected individuals.

We model the problem as a discrete SIR epidemic model, defined by parameters *G* (graph network topology), *p* ∈ (0, 1] (infection probability / speed), and *b* ∈ {0, 1, 2, 3, …} (vaccination budget per stage). We consider a network, represented by an undirected graph *G* = (*V, E*), where *V* denotes the set of nodes and *E* denotes the set of edges. Two nodes are said to be neighbors if (*u, v*) ∈ *E*. We use the notation *u* ↔ *v* to denote neighboring nodes. We use *n* to denote the number of nodes in *G*, and do not restrict ourselves to finite graphs.

Assume a spreading infection on *G* to be a discrete time contact process. An infected node infects each of its healthy neighbors with probability *p*. The state of the infection at time *t* is defined by the pair *s*_*t*_ = (*I*_*t*_, *B*_*t*_), where *I*_*t*_ and *B*_*t*_ denote the set of infected and vaccinated nodes at time *t*, respectively. At all times, *I*_*t*_ ∩ *B*_*t*_ = ∅ (i.e., a node can either be healthy, infected, or vaccinated).

The process *s*_*t*_ is initialized at some given state *s*_0_ = (*I*_0_, *B*_0_) with transitions occurring independently according to the following dynamics. If a node *i* is infected, it remains infected forever. This can be thought of as an epidemic in which all infected individuals eventually die. If a node *i* is vaccinated, it remains vaccinated forever. If a node *i* is healthy at time *t*, then node *i* stays healthy at time *t* + 1 with probability *q*_*i,t*_, where 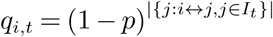. Informally, the first two conditions mean that nodes remain in their infected or vaccinated states at all times. The last condition means that at each iteration nodes infect each of their healthy neighbors independently with probability *p*.

A stationary control policy *π*(*s*) ∈ {0, 1}^*V*^ determines at each state *s* = (*I, B*) what set of healthy nodes to vaccinate. Once a set of nodes is chosen to be vaccinated they are added to *B*. We impose a budget constraint of the form

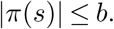

### 2.1. Criterion for Containment

We define an optimal policy as one which minimizes the expected number of infected individuals. Formally, for an initial state *s*_0_ = (*I*_0_, *B*_0_), the expected loss of a policy *π* given a budget *b* is defined by

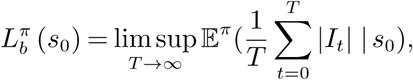

where 𝔼^*π*^ denotes the expected value induced by the vaccination policy. The optimal loss is then given by

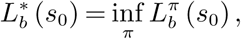

When it exists, we define the optimal vaccination policy *π*^∗^ by

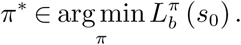

We are now ready to define our notion of containment. Given a maximal number of infected individuals (i.e., a worst-case “acceptable” loss), we ask, is a budget *b* sufficient to ensure an infection does not grow to be larger than some given value? Specifically, given a threshold *θ* ∈ ℕ ∪ {∞}, we wish to find a minimal budget such that 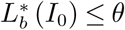 when *θ <* ∞, or 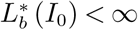 when *θ* = ∞. We define containment formally below.

#### Definition 2.1.

*Let θ* ∈ ℕ ∪ {∞}. *Let G* = (*V, E*) *be a graph on n nodes, and s*_0_ = (*I*_0_, *B*_0_) *be an initial state. When θ <* ∞, *we say that* <u>*b contains I*_0_ *in θ*</u> *if* 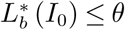. *When θ* = ∞, *we say that <u>b contains I</u>*<u>_0_</u> *if* 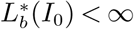.

#### Definition 2.2.

*We say that b*_*θ*_ *is a <u>weak upper bound</u> if for any b > b*_*θ*_, *b contains I*_0_ *in θ. We say that b*_*θ*_ *is a <u>weak lower bound</u> if for any b < b*_*θ*_, *b does not contain I*_0_ *in θ. We say b*_*θ*_ *is a <u>tight bound</u> if b*_*θ*_ *is both a weak upper and lower bound. We define equivalently for the case of θ* = ∞ *and denote the containment bound by b*_∞_.

## 3. Preliminaries

We use the notation [*z*]_+_ for any *z* ∈ ℝ to denote the positive part of *z*. We denote the set of edges in a graph by *E* and use the notation *u* ↔ *v* to denote (*u, v*) ∈ *E*. We denote by Δ the maximal node degree in a given graph. The number of neighbors of a set of nodes is defined to be the number of connected nodes to that set, and is denoted by *N* (*A*) for some *A* ⊂ *V*. In a similar way, for a state *s* = (*I, B*), its neighborhood is defined as the set of nodes *N* (*s*) = *N* (*I*)\*B*.

Another important measure for sets is their cuts. A cut of a set of nodes in a graph is defined to be the number of edges exiting the set. For any two sets *A, B* and a node *v*, the cut from *A* to *v* is defined by cut (*A, v*) = |{(*u, v*) ∈ *E* : *u* ∈ *A*}|. The cut from *A* to *B* is then defined by cut (*A, B*) = ∑_*v*∈*B*_ cut (*A, v*). We can then define the cut of *A* as the cut from *A* to *N* (*A*), that is, cut (*A*) = cut (*A, N* (*A*)). Equivalently, for a state *s* = (*I, B*) and node *v* ∈ *N* (*s*), the cut from *s* to *v* is defined by

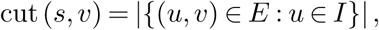

and the cut of *s* by cut(*s*) = cut (*I, N* (*s*)) = Σ_*v*∈*N* (*s*)_ cut (*s, v*).

## 4. Growth Rate

This section develops the required machinery for developing upper and lower bounds on the optimal loss. These bounds enable us to answer questions regarding containment objective. We define the notion of the growth rate. Informally, the growth rate of an infection at time *t* is the expected cardinality growth of *I*_*t*_ between time *t* and *t* + 1. That is, if we denote by *GR*_*t*_ the growth rate at time *t*, then

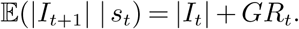

The growth rate thus changes at each time step *t*. Intuitively, a good policy would (1) maintain a small growth rate throughout the propagation of the infection and (2) bring the growth rate to zero as quickly as possible. We define the notion of the growth rate formally below.

### Definition 4.1.

*The growth rate of a set A to a set B for infection speed p is defined by*

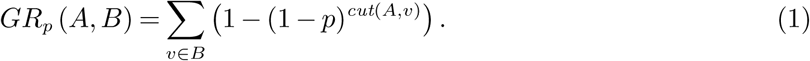

*The growth rate of a set A is defined using equation (1) as GR*_*p*_ (*A*) = *GR*_*p*_ (*A, N* (*A*)). *Equivalently, we define the growth rate of a state s* = (*I, B*) *to be GR*_*p*_ (*s*) = *GR*_*p*_ (*I, N* (*s*)).

The growth rate of a state *s* = (*I, B*), *GR*_*p*_ (*s*), takes into account the vaccinated set of nodes *B*. It can be useful to write it using the growth rate for sets in the following way:

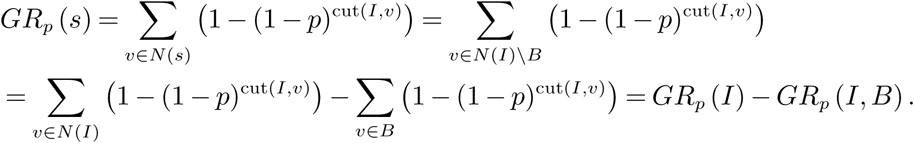

The notion of growth rates takes into account both the topological structure as well as the ever changing set of vaccinated individuals. It is thus a key characteristic of a spreading epidemic. Due to its complex nature, we look for ways to approximate it, as discussed next.

### 4.1. Maximal and Minimal Growth Rates over Upward Crusades

Growth rates give us a tool for measuring the expected growth of an infection at a time *t*. We consider the full propagation of the epidemic using the notion of maximal and minimal growth rates. These tell us how large or how small the growth rate can be under a specific policy in a given time window. This is done by considering all possible future scenarios of infection and vaccination under a given policy. We then find representative worst, best, and average case scenarios. Formally, we start by defining upward crusades as sequences of possible states under a given policy. We then define the maximal and minimal growth rates as the worst and best case growth rates over all possible sequences.

An upward crusade is a sequence of infection states which follow all possible realizations of infections starting at some initial state *s*_0_ under some policy *π*.

#### Definition 4.2.

*For a state s* = (*I, B*) *and policy π, an upward crusade of length k is a sequence u* = (*s*_0_, …, *s*_*k*_) *of k* + 1 *pairs (states), s*_*i*_ = (*I*_*i*_, *B*_*i*_), *with the following properties:*

1. *s*_0_ = *s*
2. *I*_*i*_\*I*_*i*−1_ ⊆ *N* (*I*_*i*−1_)\*B*_*i*_, *for i* = 1 … *k*
3. *B*_*i*_\*B*_*i*−1_ = *π*(*s*_*i*−1_), *for i* = 1 … *k*

*We denote the final state of the sequence by u*_*k*_. *We also denote the set of all upward crusades of length k initiating at a state s by* 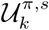. *Figure 1 depicts examples of upward crusades on a simple graph*.

**Figure 1.**
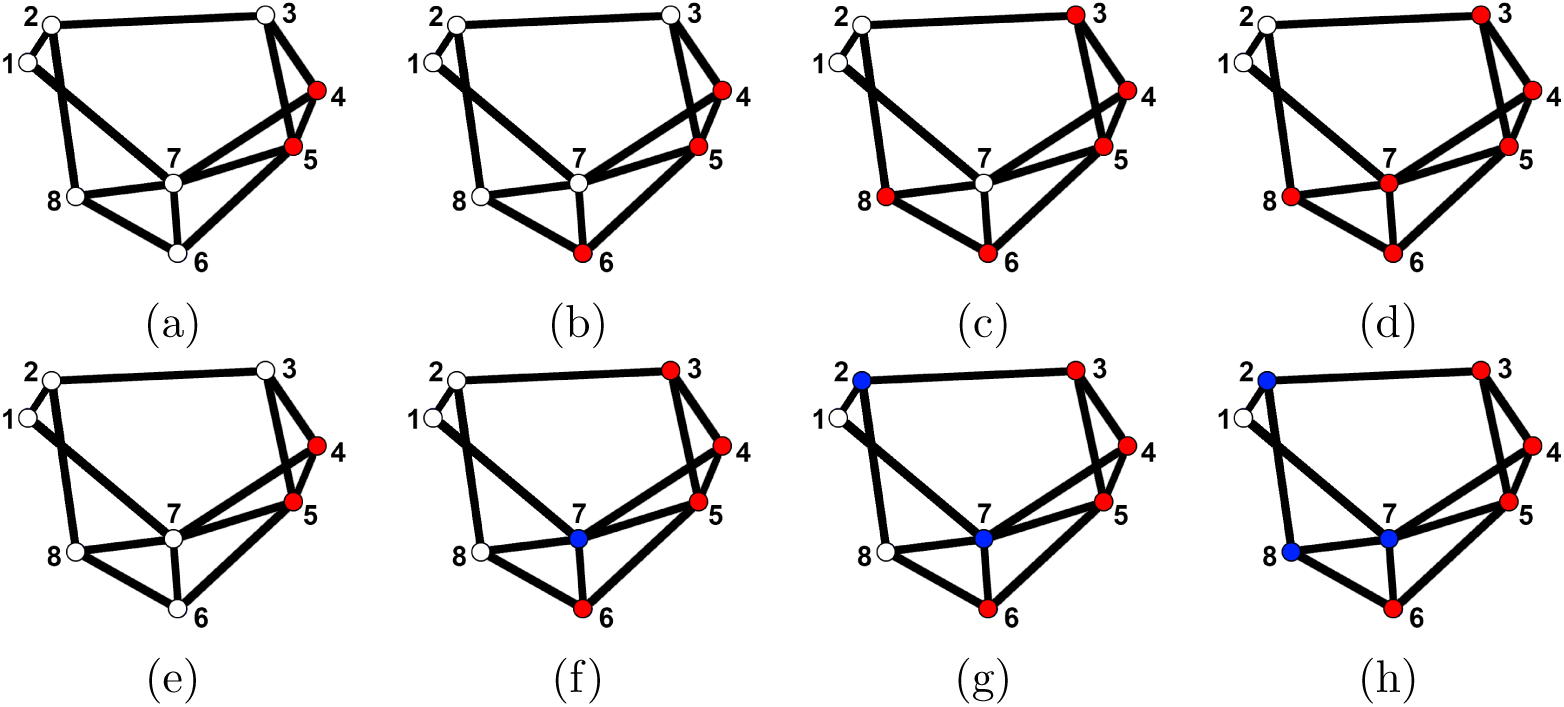
(a-d) An example of an upward crusade 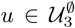, where *I*_0_ = {4, 5}. The sequence depicted is *u* = (({4, 5}, ∅), ({4, 5, 6}, ∅), ({3, 4, 5, 6, 8}, ∅), ({3, 4, 5, 6, 7, 8}, ∅)). Throughout the whole sequence, *b* = 0 and *Bt* = ∅. (e-h) An example of an upward crusade 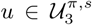 of some policy *π*Π under a budget of *b* = 1, and *s* = (*I*, ∅), where *I* = {4, 5}. The sequence depicted is *u* = (({4, 5}, {∅}), ({3, 4, 5, 6}, {7}), ({3, 4, 5, 6}, {2, 7}), ({3, 4, 5, 6}, {2, 7, 8})). Note that between states *s*2 and *s*3, the infected set does not change.

**Figure 2.**
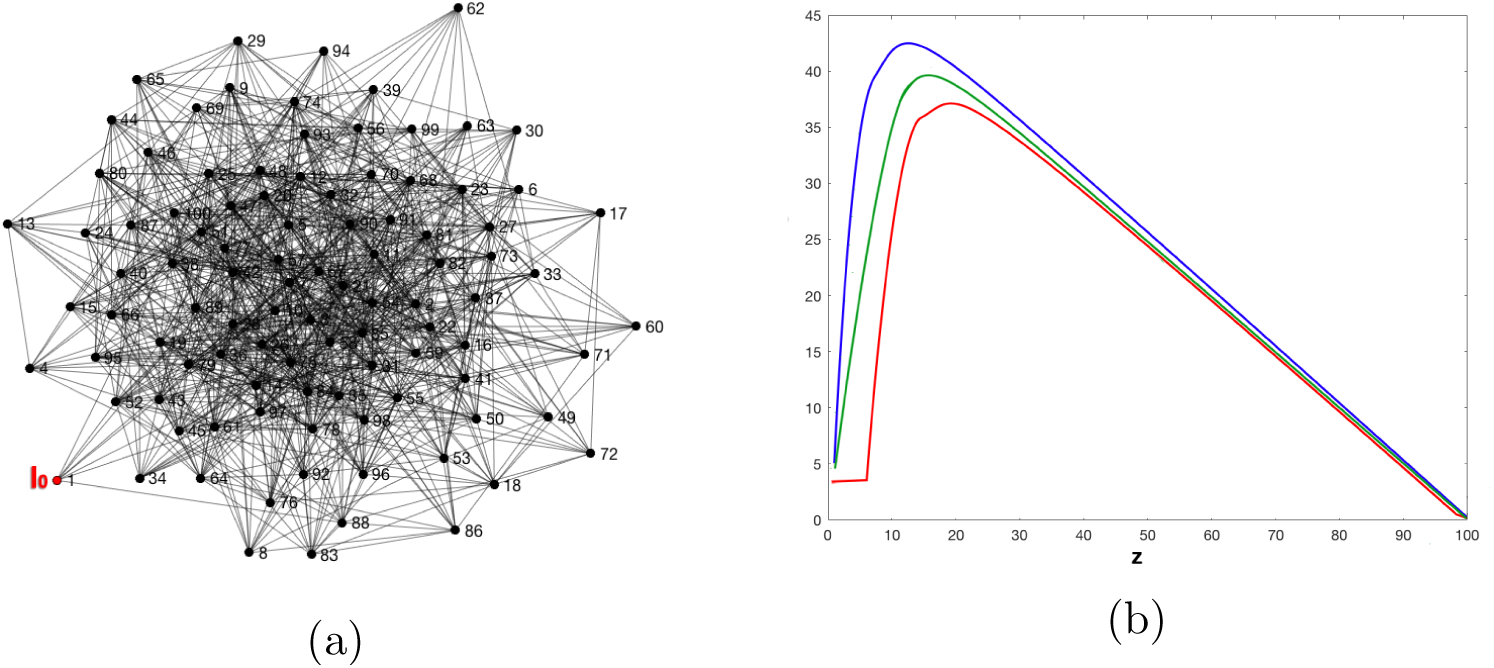
(a) A sample graph of an Erdős-Rényi model *G*(*n, s*), with parameters *n* = 100, *s* = 0.1 with diameter 3. (b) Plots of 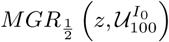 (in blue), 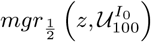 (in red), and 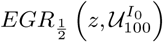 (in green) as functions of *z*, with *I*0 = {1}. Estimates were made using Monte Carlo simulations (see Section 6 for details).

Indeed, upward crusades consider all possible futures of a spreading epidemic given a vaccination policy *π*. Following the definition of upward crusades, the maximal growth rate looks *k* steps into the future, until the point when an infection of cardinality *c* is reached under some policy *π*. It then returns the worst case growth rate over all corresponding upward crusades. A similar idea follows for the minimal and expected growth rates, as defined formally bellow.

#### Definition 4.3

(Maximal and Minimal Growth Rates). *Given a policy π, a state s, and the set of upward crusades of length k*, 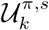, *the maximal and minimal growth rates are functions MGR, mgr* : 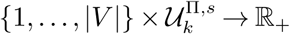, *defined by*

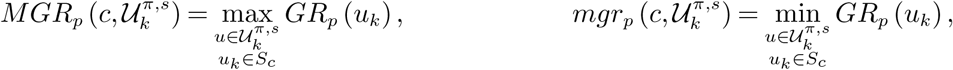

*where S*_*c*_ = {*s* = (*I, B*) : |*I*| = *c*}, *and recall that u*_*k*_ *denotes the final state in an upward crusade u. In other words, the maximum / minimum growth rates maximize the growth rate over all states that end upward crusades in* 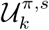 *and have infection cardinality c*.

#### Definition 4.4

(Expected Growth Rate). *Given a policy π*Π, *a state s, and the set of upward crusades of length k*, 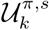, *the expected growth rate is a function* 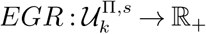, *defined by*

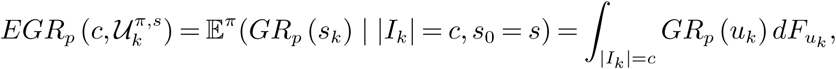

*where u*_*k*_ = (*I*_*k*_, *B*_*k*_) *is the random variable of the final state in upward crusades of* 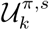, *such that the integral is taken over final states with cardinality* |*I*_*k*_| = *c*.

It is helpful to consider maximal, minimal, and expected growth rates of an empty policy which does not vaccinate any nodes. We will denote these using upward crusades 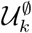 in place of 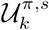 to emphasize the fact the maximum/minimum/expectation is taken over crusades which do not vaccinate any nodes (i.e., *b* = 0).

## 5. Containment

We are now ready to state our main result - explicit bounds on containment of epidemics. In order to obtain these bounds, we construct upper and lower bounds on *MGR* and *mgr*, as defined in the previous section. These allow us to calculate explicit upper and lower bounds for containment. Given an initial state of infection *s*_0_ = (*I*_0_, ∅) we denote function *l*_*α,β*_ for some parameters *α, β* ∈ ℝ_+_ (which may depend on *p*) as

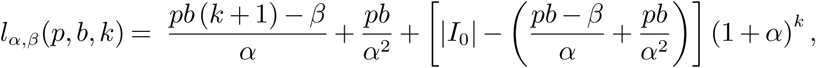

and let *k*_*b,p*_ be defined by

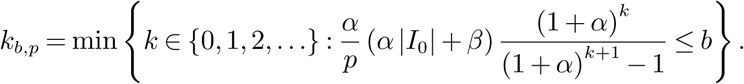

These two functions are related to the number of expected infected individuals (*l*_*α,β*_(*p, b, k*)) and the expected time for containment (*k*_*b,p*_). They are used by following theorem, which lets us calculate explicit containment bounds. We provide conditions for containing an infection in expectation whenever *MGR* and *mgr* can be bounded by linear functions. For brevity, we use the notation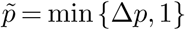, where Δ is the maximal degree in *G*.

### Theorem 5.1

(Containment Bounds). *Suppose* ∃*α, β, γ, δ*∈ ℝ _+_ *for which*

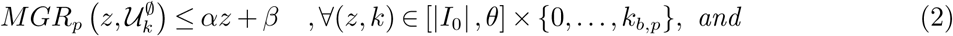

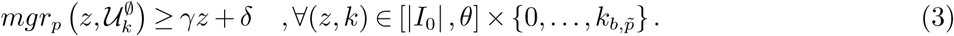

*Then*

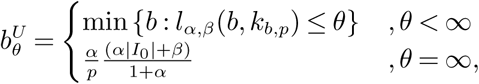

*And*

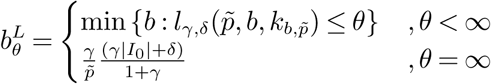

*are a weak upper and lower bounds for containment, respectively. Moreover there exists a vaccination policy π for which*,

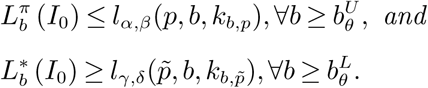

### Corollary 5.1.

*Suppose* ∃*α, β* ∈ ℝ_+_ *for which*

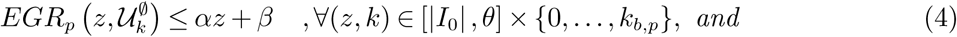

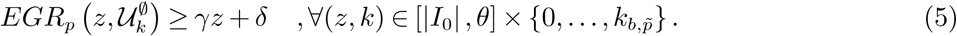

*Then all of the results of Theorem 5.1 hold*.

Note that the above theorem determines explicit upper and lower bounds for the budget needed to contain an infection. The gap between 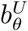 and 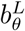 implies the error induced by the need to approximate the growth rate statistics through affine upper and lower bounds. The lower this gap, the more strict these bounds are, making the budget a necessary and sufficient condition for containing an epidemic.

### 5.1. Obtaining Explicit Bounds

We look for explicit bounds in three regimes, including d-regular trees, d-dimensional grids, and Erdős-Rényi random graphs graphs. While grids provide local connectivity, they lack long-range connections. Grids characterize slow spreading epidemics with small neighborhood growth, through contact networks that are very correlated with geography. Erdős-Rényi random graphs, on the other hand, model the opposite extreme, in which all nodes are uniformly connected, modeling large neighborhood contact networks. Finally, trees model exponential neighborhood growth. While tree contact networks are rare, spreading epidemics are often locally tree-like.

We turn to find explicit bounds on these topologies. To achieve these bounds, we apply Theorem on linear upper bounds for 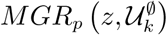, and linear lower bounds for 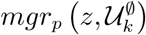. Exceptional is the case of regular trees, where the maximal and minimal growth rates coincide with a linear function, giving us tight bounds for containment.

#### 5.1.1. D-Regular Trees The following tree gives a tight bound on *mgr, MGR* on regular trees

##### Theorem 5.2.

*Let T be a regular tree of degree d. Suppose I*_0_ *is a connected set such that root* ∈ *I*_0_. *Then*

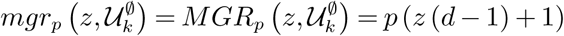

Theorem 5.2 states that whenever *root* ∈ *I*_0_, the growth rate is equal to the maximal and minimal growth rates. Applying Theorem 5.1 for the case of *root* ∈ *I*_0_, we obtain a *tight* bound for containment. Specifically, letting *α* = *p*(*d* − 1) and *β* = *p*, then

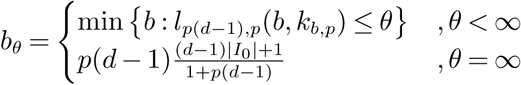

is a *tight* containment bound. Moreover, we can obtain the exact number of expected infected individuals under such (optimal) vaccination strategy. Specifically, the number of expected individuals is given by

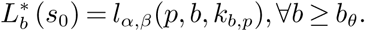

Notice the special case of *I*_0_ = *root* and *θ* = ∞ where the *tight* containment bound becomes

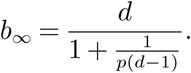

This result is non-trivial in the sense that as *p* decreases, a budget lower than *d* (the degree of the tree) is necessary in order to contain an infection initiated at the root. That is, when *p ≠* 1, even if an infection cannot be stopped at its initial state, it can still be contained at a future state.

#### 5.1.2. D-Dimensional Grid

In a conjecture made in [15], a minimal budget *b* = 2*d* − 1 is needed to contain a deterministic contact process starting at a single node on the *d*-dimensional grid. This conjecture was later proven in [16]. In this section we prove a more general result. Specifically, we obtain the minimal budget needed to contain an epidemic on the *d*-dimensional grid, as provided by the following theorem.

##### Theorem 5.3.

*Let I*_0_ *be a connected set of initially infected nodes on the d-dimensional grid. We have that*

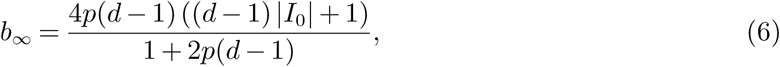

*is a weak upper bound for containment*.

Notice the special case of |*I*_0_| = 1 and *p* = 1. In this case we get that

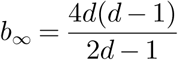

is a weak upper bound for containment, which equivalently means an infection can be contained for any *b* which satisfies

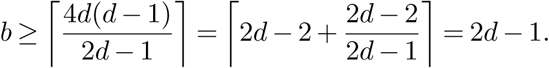

That is, Equation (6) proves the bound proven in [16] for a more general case of *p <* 1.

Next, we look for weak lower bounds for containment. In order to use Theorem 5.1, we must lower bound *mgr* by a linear function. Unfortunately, on the *d*-dimensional grid, *mgr* is a concave, non-monotonic function. One method to overcome this problem is to lower-bound *mgr* by a piecewise linear function, enabling us to apply Theorem 5.1 recursively on finite intervals. Lemma 5.1 provides such an affine, monotonically non-decreasing lower bound for *mgr* on the *d*-dimensional grid on a finite interval.

##### Lemma 5.1.

*Let G* = (*V, E*) *be a d-dimensional grid, and let a, θ* ∈ N *with a < θ. Then*

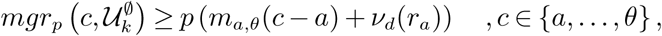

*Where*

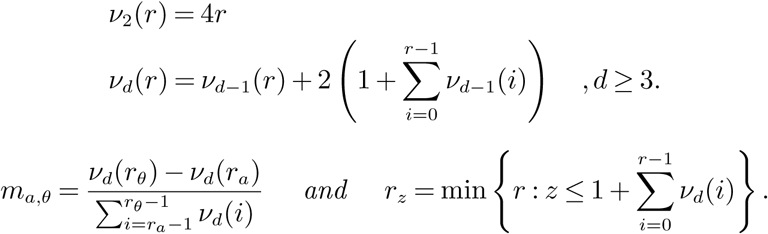

One can thus utilize Lemma 5.1 to estimate a weak lower bounds for containment on the *d*- dimensional grid.

#### 5.1.3. Erdős-Rényi We finally consider Erdős-Rényi random graphs

Let *G*_*n,s*_ be an Erdős-Rényi model, where *n* denotes the number of nodes, and *s* the edge sampling probability. We have the following result.

##### Theorem 5.4.

*Given an Erdős-Rényi model G*_*n,s*_ *with* 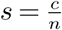 *for some constant c >* 1,

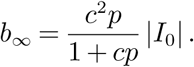

*is a weak upper bound for containment*.

This containment bound is mainly due to the expected cut under an expected initialization of the graph. We can view this result at two extreme cases. When *cp* ≫ 1 the bound becomes *b*_∞_ = *c* |*I*_0_|. This models very clustered networks with very quickly spreading epidemics. This condition can be thought of as a worst case spreading epidemic. On the contrary, when *cp* ≪ 1 the bound becomes *b*_∞_ = *c*^2^*p* |*I*_0_|. This condition models a slowly spreading epidemic on a sparse contact network. This can be thought of as a best-case scenario of a spreading epidemic. Indeed, a small vaccination budget is needed to ensure containment.

Similar to the *d*-dimensional grid, the expected growth rate of an Erdős-Rényi graph is concave. We therefore lower bound the expected growth rate by a piece wise linear function, as stated below.

##### Proposition 5.1.

*Let G*_*n,s*_ *be an Erdős-Rényi model. Then*

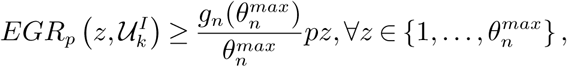

*where g* : N → ℝ_+_ *is defined by* 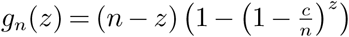 *and* 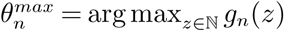

Proposition 5.1 gives us a method to compute a lower bound on *EGR* for Erdős-Rényi random graphs. Applying Corollary 5.1 would thus provide us with lower bounds for containment.

## 6. State-Dependent Budget

In the previous section we have shown how Theorem 5.1 can be applied to well known toplogies that have been studied extensively in the literature. In this section we show how Theorem 5.1 can be used to construct a state-dependent budget allocation policy, which suggests a specific budget for every time step, according to the state’s current *EGR*/*mgr* characteristic. We show this strategy acheives better containment on two real world networks, when compared to a constant budget strategy which consumes an equal global budget.

Recall Theorem 5.1 gives guarantees for containtment of an infection when *mgr* can be lower bounded by a linear function under a minimal budget *b*^*L*^. In fact, when only a lower bound in

[|*I*_0_|, *θ*]] is given, then 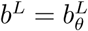 is a lower bound for containment in *θ* (i.e., the infection can be contained in expectation with cardinality at most *θ*). In theory, given *mgr/EGR*, a controller can choose at each state *s*_*t*_ = (*I*_*t*_, *B*_*t*_) an objective *θ*_*t*_ = |*I*_*t*_| + *M*_*t*_, for some *M*_*t*_ ∈ N. Then, by lower bounding *mgr/EGR* in [|*I*_*t*_|, *θ*_*t*_] and applying Theorem 5.1, a specific budget can be allocated for that state. This process can be repeated at each time step in order to obtain a “minimal” budget according to the current lower bound in [|*I*_*t*_|, *θ*_*t*_].

In contrast to trees and grids (as well as other symmetric topologies), an explicit expression of *mgr/EGR* is hard to obtain. Even a lower bound may be complicated to express. For this reason, Monte Carlo simulations can be used in order to approximate *mgr/EGR*. Specifically, let *τ* = {*u*_1_, *u*_2_, …, *u*_*T*_} be *T* trajectories of length *d* starting at state *s*. We can approximate a lower bound on *mgr* for *d* steps by

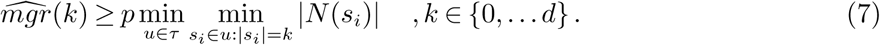

Similarly, a lower bound on *EGR* can be approximated by

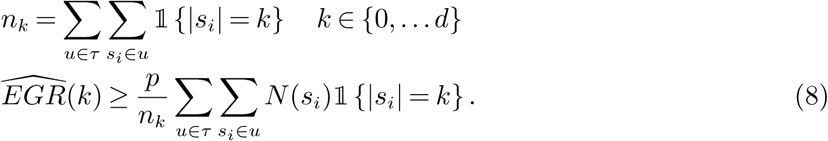

Using the lower bounds in Equations (7) and (8) a simple linear lower bound can be constructed. Finally, Theorem 5.1 can be applied to these linear lower bounds. Algorithm 1 gives a state-dependent budget allocation procedure which applies Theorem 5.1 with an approximated linear lower bound to *mgr/EGR* using Monte Carlo simulation.

### 6.0.1. Experiments

We have run our simulations on two real email networks: the Enron email communication network [17], which covers all the email communication within a dataset of approximately half million emails, and the EU email communication network [18], generated using email data from a large European research institution for a period of 18 months. In part, using email networks is motivated by the fact that many computer viruses spread by email attachments. We have chosen to test our algorithm on these networks due to their broad degree distributions. In our simulations we have used the a first-acquaintance policy which attempts to minimize the growth rate by minimizing the cut of the current infection, i.e., *π*^*CUT*^ (*s*) ∈ arg min {cut (*s*)}.

#### Algorithm 1: State Dependent Budget Allocation

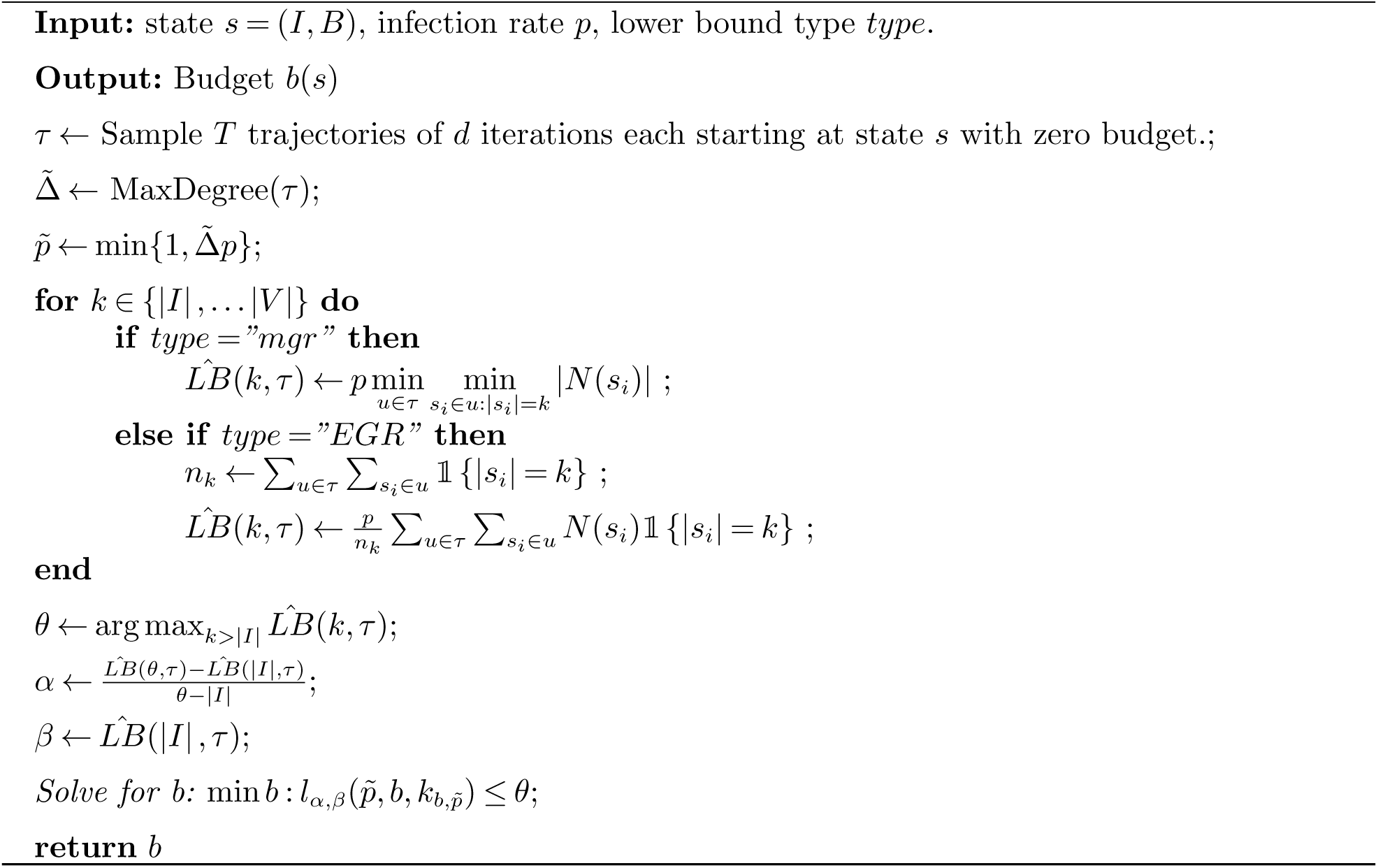

Results of our simulations are plotted in Figure 3. Statistical data on the two networks in provided in Table 1. In all of our experiments we picked starting nodes uniformly at random ^1^. We averaged the results over 100 uniform samples of initial states. We used initial cardinalities of 2000 for the Enron network and 5000 for the EU network. Each sample was run 30 times. We’ve tested lower bounds on *EGR* and *mgr* using Monte Carlo simulations of length 3. All our experiments used an infection probability of *p* = 0.05.

**Table 1.** Dataset statistics of the Enron and EU email networks.

|  | Nodes | Edges | Nodes in<br>largest<br>WWC | Nodes in<br>largest<br>SSC | Average<br>clustering<br>coefficient | Number of<br>triangles | Diameter |
| --- | --- | --- | --- | --- | --- | --- | --- |
| Enron | 36692 | 183831 | 33696 | 33696 | 0.4970 | 727044 | 11 |
| EU | 265214 | 420045 | 224832 | 34203 | 0.0671 | 267313 | 14 |

**Figure 3.**
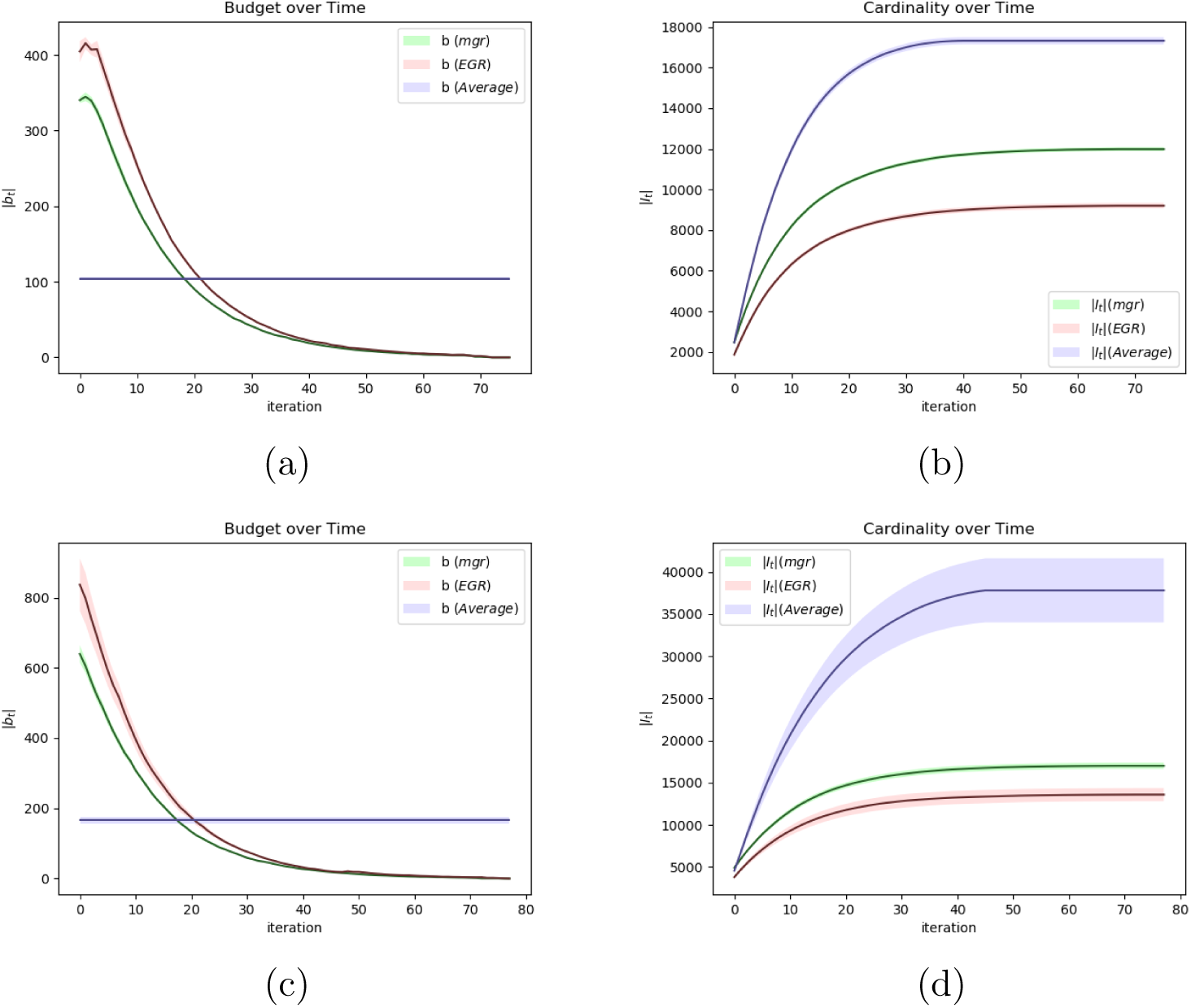
Plots comparing different budget allocation strategies on the Enron email network of 36,692 nodes (Plots a,b) and EU research institution email network of 265,214 nodes (Plots c,d). Plots show state dependent budget allocation using *mgr* (green), *EGR* (red), and constant budget equal to the average budget used by *EGR* (blue). Initial cardinalities that were tested are 2000 infected nodes for Enron network, and 5000 infected nodes for EU network. Infection rate was *p* = 0.05 for both networks. Both networks were sampled 100 times and averages over 30 runs per sample. Light areas show standard deviation from mean.

We compared our results to a constant budget allocation strategy. The constant budget was calculated by taking the maximal total budget used by our algorithm and dividing by the average number of iterations used. Specifically, denote by 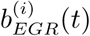 and 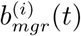 the budget used in test *i* by the *EGR* and *mgr* lower approximations, respectively. Also, denote by *T*_*avg*_ the average number of iterations for containment under these approximations. Then,

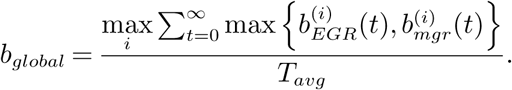

Results, as depicted in Figure 3, clearly show that a state dependent budget strategy which allocates more resources at the beginning of an infection is superior over one that only uses a constant budget with an equal global budget.

## 7. Conclusion and Implications for the COVID-19 Outbreak

During the pandemic outbreak of COVID-19 in 2020, governments all over the world announced movement restrictions which included remaining at home, reducing public transport, closure of places of worship, schools, commercial/dining establishments, heritage sites, and parks/beaches. Such restrictions altered the population connectivity graph’s topology, reducing connectivity, and allowed for better control of the spreading epidemic. Nevertheless, such extreme measures threaten to devastate economies and ramp up inequality. These losses would reverberate across societies, impacting education, human rights and, in the most severe cases, basic food security and nutrition. This work is focused on the question: Can containment be guaranteed in expectation with a given vaccination budget? And conversely, when is the spread of an epidemic inevitable? We obtained bounds for containment. These bounds tell us the minimal vaccination budget needed to ensure containment of an epidemic, as well as the minimal budget under which an outbreak would occur. In Section 6 we constructed an algorithm that uses lower bounds on *mgr/EGR* for state dependent budget allocation. We tested this algorithm using a first acquaintance vaccination policy suggesting such budget allocation strategy outperforms constant budget allocation with an equal global budget.

It is clear from our results that the graph’s topology has a tremendous effect on the number of infected nodes as well as our ability to contain it. D-dimensional grids represents contact graphs in which an infection can spread by spatial proximity. They model infection spreads such as a Bluetooth virus or human sickness. On the other hand, Erdős-Rényi random graphs form networks with low diameter. This topology models an infection spreading over long distance, such as the Internet or over most “normal” social networks. We provide a method for relating the topology of the graph to the minimal budget needed for containment. We can assess the average degree needed to manage an epidemic under limited vaccination resources using an Erdős-Rényi model or a grid model. This insight may allow governments to mitigate Draconian closure laws that could have calamitous effects on society.

Our results suggest that the concept of “herd immunity”, advocated by some, depends critically on the degree of the connectedness of the society. If the ties of the society are weak enough, then immunity by a relatively small fraction of society would suffice for the epidemic to die out. Our containment bounds are also beneficial for preserving resources including the number of needed vaccines, the rate in which they must be administered, as well as required personnel for administering vaccines. Assuming governments have a certain capacity of controlling infected individuals, our upper bounds for containment may give the needed insight for saving resources, while preventing hospital overloads, ventilator shortage, etc.

## Data Availability

All data used is open source

## Footnotes

1 Choosing sources in a realistic way is an open problem - the data that could offer a solution to this problem seems to be extremely scarce at this time.

